# Applying Behavioral Nudges in a Dietary Comparator for Surgical Trials: Developing the MediDiet

**DOI:** 10.1101/2021.10.14.21265033

**Authors:** Irene Y. Zhang, Daniel Norwitz, Adam Drewnowski, Nidhi Agrawal, David R. Flum, Joshua M. Liao

## Abstract

**Introduction:** The Mediterranean diet is being studied as an alternative to surgery for common gastrointestinal conditions. However, dietary change can be challenging. “Nudges” – behavioral science strategies seeking to influence cognitive processes to promote good behaviors – may help. We conducted a pilot study evaluating the acceptability of the MediDiet, a behaviorally designed dietary intervention incorporating nudges and recommendations based on the Mediterranean Diet.

**Methods:** We conducted a three-phase pilot study involving parallel randomized surveys of U.S. adults. After completing a validated questionnaire assessing dietary consistency with a Mediterranean diet, participants were randomized to feedback containing no nudge versus nudge: peer comparison; peer comparison + positive affect induction; or default. Participants then rated their positive and negative emotions, motivation for dietary change, and interest in recipes. Responses were analyzed using baseline covariate-adjusted regression.

**Results:** Among 1,709 participants, 56% were men, 73% were White, and the mean age was 38. In response to dietary feedback, participants reported low negative emotions, high positive emotions, moderate motivation for dietary change and high interest in recipes. Nudges did not affect the extent of negative (p=0.104) or positive (p=0.34) emotions, motivation (p=0.139), or interest (p=0.86). In exploratory analyses, those with moderate and high consistency with the Mediterranean diet, with or without nudges, reported lower negative affect, greater positive affect, greater motivation, and greater interest in recipes, than the minimally consistent, no nudge group.

**Conclusion:** Delivering dietary feedback based on the Mediterranean diet using behavioral nudges was acceptable among U.S. adults, rousing positive reactions without triggering negative ones. As early evidence, this pilot study provides the basis for testing nudge-based dietary guidance among individuals with symptomatic gallstones, diverticulitis, and other gastrointestinal diseases.

**Highlights:**

- The MediDiet is being developed as a novel, behaviorally designed dietary intervention for the non-operative management of common gastrointestinal conditions including symptomatic gallstones and diverticulitis, classically managed with surgery.
- In this exploratory pilot study, delivering dietary feedback based on the Mediterranean diet using behavioral nudges was acceptable among U.S. adults.
- This foundational work will help serve as the basis for testing nudge-based dietary guidance among individuals with symptomatic gallstones, diverticulitis, and other gastrointestinal diseases, and ultimately conducting clinical trials to compare dietary interventions to surgical management for these conditions.

## Introduction

Many medical conditions that can be managed surgically also have non-operative alternatives, presenting important choices for patients. These non-surgical options often require patients to make and sustain diet and lifestyle behavior changes. For example, two of the most common gastrointestinal conditions with surgical treatments – symptomatic gallstones and diverticulitis – can be managed non-operatively with dietary change focused on fatty food avoidance and high fiber intake.

Due to well-known challenges with dietary change and inadequacy of general guidance about foods, specific dietary interventions are likely needed to maximize the benefit of non-operative management options. In particular, the Mediterranean diet (low animal fat and high fiber from legumes, nuts and seeds, vegetables and fruit), which has been demonstrated to be beneficial in randomized trials of cardiovascular disease, is now being studied for the management of symptomatic gallstones, diverticulitis, and other gastrointestinal diseases.^1–5^

The opportunity is ripe for clinical studies that compare the Mediterranean diet to surgery for the management of common gastrointestinal conditions. However, even evidence-based recommendations and counseling about the Mediterranean diet may have limited impact on patient outcomes if they do not incorporate insights from behavioral science. Diet change is fundamentally behavioral, and diet-based management options may be more effective if guidance is paired with behavioral “nudges” – strategies that acknowledge how humans process and respond to information, and seek to influence those cognitive processes to promote good behaviors.^6–8^ Nudges have been used in a number of health settings, and have shown promise including in increasing healthy dietary and nutritional choices for overweight individuals.^9–12^

Unfortunately, little is known about if and how best to provide Mediterranean diet guidance using behavioral nudges for the purposes of non-operative disease management. To address this gap and identify dietary interventions that could be studied as alternatives to surgical treatment, we conducted a multi-phase pilot study evaluating the acceptability of the MediDiet – a behaviorally designed dietary intervention that uses nudges to deliver recommendations based on the Mediterranean Diet.

## Methods

We conducted a three-phase pilot study involving parallel, two-arm randomized surveys of U.S. adults. The study was deemed exempt by the University of Washington Institutional Review Board, and informed consent was waived.

### Nudges

Given the dearth of existing knowledge on this topic and the exploratory nature of this study, we sought to evaluate the acceptability of nudges as a class of strategies, compared to no nudge. We chose three types of nudges based on their application to dietary change and other health care choices^12^: *peer comparisons* that seek to motivate behavior by providing individuals with information about their behaviors compared to those of their peers (e.g., “your diet is more consistent with a Mediterranean diet than nearly all U.S. adults”); *positive affect induction* that seeks to drive behavior by encouraging a positive, self-efficacious frame of mind (e.g., “think of a time when you felt good about your diet”); and *defaults* that prompt behavior by leveraging the tendency for individuals to choose pre-selected options (e.g., “you will be emailed 5 easy, delicious recipes based on a Mediterranean diet that you can use to improve your health; you may opt out if you wish”). Nudges were incorporated into messages that were provided to participants with feedback about how consistent their diets were with the Mediterranean diet.

### Study Population

Participants were U.S. adults (ages ≥ 18) recruited from Amazon Mechanical Turk (MTurk), a web-based crowdsourcing platform that has been used for survey-based clinical and health services research.^13–15^ Consistent with prior work, we required participants to have a history of completing surveys and excluded those with duplicate IP addresses, suspicious geocode locations, or responses signaling automated responses.^13,16^

### Survey Instrument and Outcomes

The survey was designed and administrated through the Qualtrics survey platform.^17^ Participants were asked to complete a dietary questionnaire, the Mediterranean Diet Adherence Score (MEDAS), a validated instrument that has been used in clinical trials to measure and track the consistency of one’s diet with a Mediterranean diet, on a scale of 0 (inconsistent) to 14 (most consistent).^1^

All participants were then given feedback based on their MEDAS about how consistent their diet was with a Mediterranean diet, categorized based on prior work as highly consistent (MEDAS 7 to 14), moderately consistent (MEDAS 3 to 6), or minimally consistent (MEDAS 0 to 2).^9,18–20^ These categories were then used to stratify randomization such that within each consistency category, participants were then randomized to no additional information (*no nudge*) versus one of several nudges: peer comparison; peer comparison + positive affect induction; or default.

All participants were then asked to rate their levels of positive and negative emotions, in questions adapted from the Positive and Negative Affect Schedule (5-point Likert; 1 = Not at all, 5 = Extremely),^21^ as well as motivation for dietary change and interest in new recipes (7-point Likert; 1 = Not at all, 7 = Extremely). The outcomes for the study were dichotomized versions of these responses: high positive affect (top quartile vs. other quartiles), low negative affect (bottom quartile vs. other quartiles), high motivation (top quartile vs. other quartiles), and high interest (top quartile vs. other quartiles).

Additionally, participants were asked to self-report sociodemographic information and prior dietary and cooking experiences. They received compensation of $1.00 for participation. The survey required five and a half minutes on average to complete.

### Data Analysis

Data were described using mean (standard deviation) for continuous variables and frequency (percentages) for categorical variables, and analyzed using baseline covariate-adjusted logistic regression.^22,23^ The regression models were adjusted for baseline characteristics identified *a priori* as potentially influencing the relationship between interventions and outcomes: age, gender, race, ethnicity, BMI, familiarity with a Mediterranean diet, cooking experience, income, and weekly spending on food.^1,24,25^

Main analyses compared individuals who received nudge versus no nudge in the overall cohort. In exploratory analyses, we compared outcomes between nudge and no nudge groups across different categories of Mediterranean dietary consistency – i.e., comparing the effect of nudges across individuals with diets that were highly versus moderately versus minimally consistent with the Mediterranean diet. We aimed to recruit 100 participants per stratified arm for the first two study phases, consistent with prior exploratory work.^13^ Based on those initial results, the third study phase was designed to have 80% power to detect a >15% difference in outcomes. Statistical tests were two-tailed with alpha = 0.05. Analyses were conducted in R, version 4.1.0.^26^

The funding source had no role in the study.

## Results

Among 1,709 participants in total, 56% were men, 73% were White, 83% were non-Hispanic, and the mean age was 38 years (Table 1). Overall, 43% had completed college or higher levels of education, while 41% reported an annual income of $50,000 or less. The average BMI was 25, based on self-reported height and weight. These factors were similarly distributed across the three study phases.

**Table 1.** Sample characteristics

| Characteristic | Overall, N =<br>1,709<sup>1</sup> |
| --- | --- |
| Age, mean (SD) | 38 (11) |
| Gender |  |
| Man | 952 (56%) |
| Woman | 748 (44%) |
| Other | 4 (0.2%) |
| Prefer not to say | 5 (0.3%) |
| Race |  |
| White | 1,242 (73%) |
| Black or African American | 264 (15%) |
| Asian | 120 (7.0%) |
| American Indian or Alaska Native | 12 (0.7%) |
| Native Hawaiian or Pacific Islander | 6 (0.4%) |
| Other or Multiracial | 56 (3.3%) |
| Prefer not to say | 9 (0.5%) |
| Ethnicity |  |
| Non-Hispanic | 1,413 (83%) |
| Hispanic | 265 (16%) |
| Prefer not to say | 31 (1.8%) |
| BMI, mean (SD) | 25 (8) |
| Education |  |
| High School or Some College | 593 (35%) |
| Completed College or Higher | 722 (42%) |
| Other | 394 (23%) |
| Annual Income |  |
| <\$25,000 | 190 (11%) |
| \$25,000-50,000 | 512 (30%) |
| \$50,001-75,000 | 482 (28%) |
| \$75,001-100,000 | 279 (16%) |
| >\$100,000 | 246 (14%) |

<sup>1</sup>Mean (SD); n (%)

At baseline, most participants indicated some familiarity with the Mediterranean diet. About half (54%) had experience with a dieting program in the past, while a minority (32%) had any dietary restriction. They reported an average weekly spending of $124 on food. On the dietary questionnaire, most participants reported baseline diets that were moderately consistent with a Mediterranean diet (mean MEDAS 6 +/- 3). The majority of participants reported dietary practices either highly or moderately consistent with a Mediterranean diet; approximately 10% of participants reported diets that were minimally consistent with a Mediterranean diet.

In response to feedback messages about their diet, participants reported low levels of negative emotions (e.g., sad, guilty, disappointed) and high levels of positive emotions (e.g., proud, inspired, and enthusiastic) overall. The use of nudges did not affect the extent of negative (p = 0.104) or positive (p = 0.34) emotions (Fig. 1).

**Figure 1.**
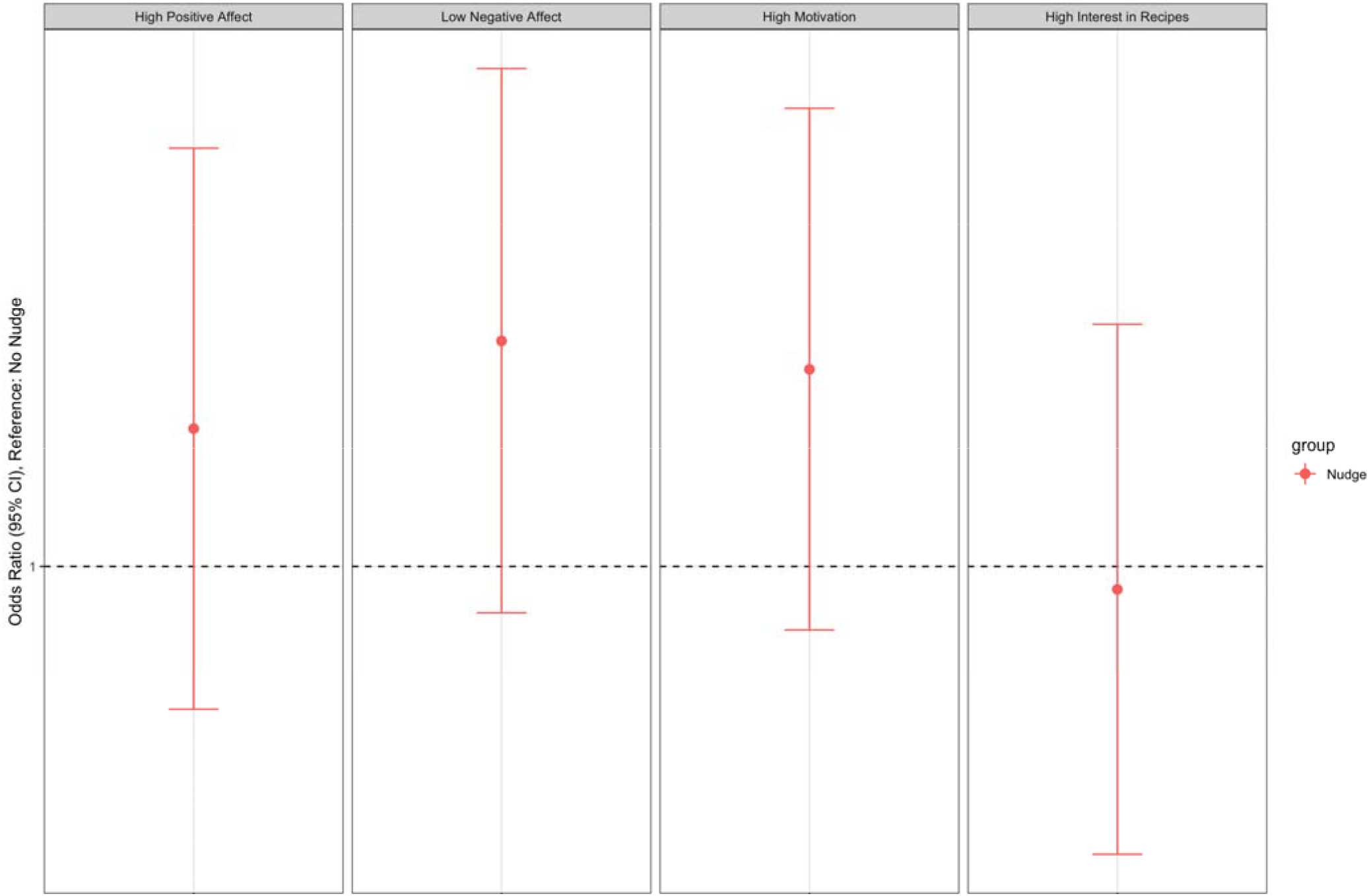
The Effect of Nudges on Study Outcomes

After receiving feedback messages, participants overall reported moderate levels of motivation for dietary change and high levels of interest in new recipes that could be used to make their diet more consistent with a Mediterranean diet. Motivation and interest in new recipes did not differ based on the use of nudges (p = 0.139 and p = 0.86, respectively) (Fig. 1).

In exploratory analyses, affect, motivation, and interest in new recipes were consistent between nudge versus no nudge groups within each dietary consistency category, but varied across categories. Compared to individuals in the minimally consistent category that did not receive a nudge, those in the moderately consistent category reported greater positive affect, lower negative affect, and greater motivation and interest in recipes whether they received a nudge or not (Fig. 2). Similarly, all participants with diets highly consistent with the Mediterranean diet reported greater positive affect, lower negative affect, and greater motivation and interest in recipes, as compared to participants in the minimally consistent category who did not receive a nudge (Fig. 2).

**Figure 2.**
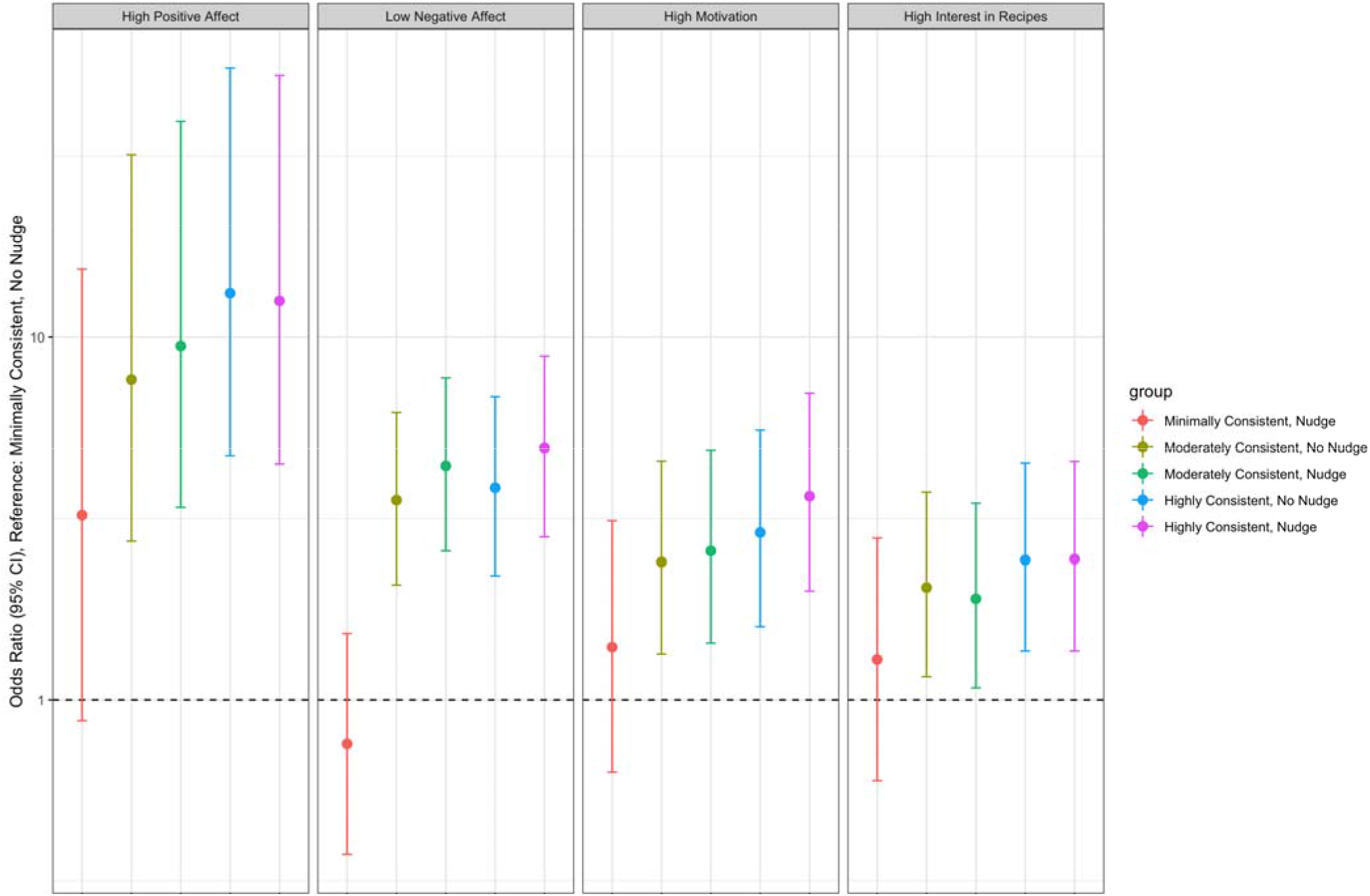
The Effect of Nudges on Study Outcomes, Stratified by Study Arm and Dietary Consistency Categories

## Discussion

In this exploratory pilot study, delivering dietary feedback based on the Mediterranean diet using behavioral nudges was acceptable among U.S. adults. This approach did not adversely affect emotions, motivation for dietary change, or interest in new recipes, overall or within strata of individuals with dietary habits that varied in consistency with the Mediterranean Diet.

This study fits into a larger body of work on dietary interventions for the purpose of disease management. Adherence to dietary interventions has been reported to be as high as 89% for the management of cardiovascular disease, but only 30% of individuals with symptomatic gallstones can manage the condition with dietary change under current practices.^27,28 29^ Methods for improving adherence in prior work have varied,^28,30,31^ and the extent to which they have been applied systematically in clinical practice is not clear. This context helps underscore the need and rationale for this study and future work designing elements of the MediDiet – to identify effective dietary interventions for disease management, and assess the potential of nudges for doing so.^12^

Study limitations include a non-clinical setting and participants that may not have the medical conditions that would ultimately be targeted by the MediDiet intervention. Most participants also reported dietary practices that were consistent with the Mediterranean diet at baseline. Additionally, three candidate nudges were selected for this exploratory study, but other types of nudges should be tested and compared directly in future work.

Nonetheless, as early evidence, this multi-phase pilot study provides foundational work that can serve as the basis for additional validation and testing. In particular, our findings help provide the basis for testing nudge-based dietary guidance among individuals with symptomatic gallstones, diverticulitis, and other gastrointestinal diseases, and ultimately conducting clinical trials to compare dietary interventions to surgical management for these conditions.

## Data Availability

All data produced in the present study are available upon reasonable request to the authors.

## Acknowledgements

Dr. Zhang was supported by the National Institute of Diabetes and Digestive and Kidney Diseases of the National Institutes of Health under Award Number T32DK070555. This work was also funded in part by a generous gift from Marty and Linda Ellison. The content is solely the responsibility of the authors and does not necessarily represent the official views of the National Institutes of Health.

## Notes

### Competing Interest Statement

Dr. Liao reports grants from the National Institute of Aging outside of this submitted work.

### Author Declarations

The study was deemed exempt by the University of Washington Institutional Review Board.

